# Contexts enhance ratings of craving and psychophysiological responses of cue-reactivity in tobacco use disorder

**DOI:** 10.1101/2022.07.12.22277347

**Authors:** Sabine Vollstädt-Klein, Frauke Nees, Alfred Wieland, Damian Karl, Carsten Diener, Michael N. Smolka, Herta Flor

## Abstract

**Introduction:** Confrontation with smoking-related cues can provoke craving and relapse, even after long periods of abstinence. The context of these cues might be important for the elicitation of craving. In this study we examined the effects of social, physical and consumptive contexts on cue-reactivity in smokers using ratings and physiological measures.

**Method:** A total of 22 smokers (12 male, 10 female) with tobacco use disorder (TUD) assessed using a cue-reactivity picture perception paradigm in a laboratory setting. Pictures of smoking in different physical and social contexts and of different phases of the smoking ritual were used. Ratings of cigarette craving, valence and arousal as well as startle reflex modulation, skin conductance responses and heart rate were assessed.

**Results:** We observed an increased startle modulation in the bar versus control context and a stronger heart rate deceleration in the social versus neutral context. Social smoking induced more craving than neutral context, and cues from the beginning of the smoking ritual induced more craving than in the terminal phase. Social smoking context was rated as more arousing than neutral context, and cigarettes of the terminal phase of the smoking ritual were rated as least pleasant.

**Conclusions:** The results indicate that physical, social and consumptive contexts differentially affect cue-reactivity in TUD.

**Implications:** Cues in contexts lead to more pronounced indicators of craving than cues alone. Thus, contexts should be included in both, the assessment and treatment of TUD.

## Introduction

Although most smokers are aware of the negative consequences of tobacco consumption, they do not quit smoking. Yet tobacco consumption as a function of its associated secondary diseases remains one of the most common causes of preventable disease worldwide[1]. Out of 111 000 deaths, 20 000 can be attributed to pulmonary disease[2] as a function of long-term tobacco consumption. The development and maintenance of tobacco use disorder (TUD) involves mechanisms related to classical conditioning, genetic predispositions, situational, emotional and motivational influences, as well as neuro- and psychobiological processes.

However, these models do no fully explain all relevant mechanisms. For example, the significance of craving in the context substance use disorder has not been completely clarified[3]. The phenomenon whereby substance-related cues are able to elicit diverse responses in individuals with SUD – so called cue reactivity – is demonstrable, but the correlation between ratings of craving and measured physiological reactions is not very high (correlation coefficients between r = 0.38 and 0.52). Verheul, van den Brink and Geerlings[4] therefore assumed that there may be different types of craving. These are likely mediated by processes involving positive reinforcement, negative reinforcement, or loss of control. Loss of control in this case refers to the inability to resist appetitive or aversive stimuli.

Classical conditioning is an important model that contributes to the understanding of mechanisms driving the maintenance of SUD, including nicotine and alcohol use disorder. Robinson and Berridge[5] postulated that through a process of learning, certain stimuli become associated with a particular drug and are then able to evoke a conditioned motivational state in an individual with SUD that promotes further drug-seeking. At the basis of this learning process are the activation of the reward system and a permanent neuroadaptation of mesolimbic dopamine signaling.

These mechanisms can be activated by nicotine-associated cues even in the absence of nicotine consumption and regardless of whether the expected reward is realized or not. As a result, pictures representing the act of smoking have the same effect as the addictive cigarette itself. However, it must be noted that in one third of all test persons evaluated in a laboratory setting, no effect of drug-related cues could be found[6, 7]. This finding could be related to variations in emotional disposition (for example, anxiety)[8], personality-specific traits, as well as SUD-related aspects (for example, severity of SUD, environmental stimuli)[9].

The effects of nicotine and nicotine-related cues are not only mediated by their psychophysiological consequences, but also depend on the particular situation and context within which the smoker finds themselves[10-13]. A functional magnetic resonance imaging (fMRI) study demonstrated that personal smoking environments influenced neural activation as well as craving[14]. Schupp, Mucha and Pauli[15] demonstrated that in a controlled field- and laboratory study, situational contexts modulated the pharmacological mechanism of actions of nicotine. They also demonstrated differential neural activation and arousal patterns for the beginning versus the terminal phase of the smoking ritual[16]. Further, they found an attentional bias to cues from the beginning of the smoking ritual, which was associated with neural activation[17]. Another predictor for developing a smoking habit is overall emotional and psychological well-being. An indirect association between smoking and psychological well-being exists[18]. In addition to internal, external cues such as social situations or physical environments associated with smoking may also modulate cue reactivity and craving[19]. Exposed to a situation containing a given stimulus, the organism reacts with a behavior composed of emotion, cognition and action[18]. In the present study, we employed the startle reflex paradigm, a psychophysiological method, as well as stimulus ratings [19] in order to measure test subjects’ emotional response to smoking cues in various contexts.

The present study investigated whether social and other typical smoking contexts increase cue reactivity in smokers in comparison to neutral and contextless situations. Valence and arousal as well as craving were measured in response to the presentation of various smoking cues with and without context. Psychophysiological responses were assessed using the electromyogram (EMG) to record startle reflex modulation, skin conductance responses, and the electrocardiograph (ECG) to record heart rate. The consumption status of a cigarette was also viewed as an important contextual influence and we thus examined how the consumption status of a cigarette might affect the degree of physiological and psychological craving produced.

We thus hypothesized that contextual stimuli would result in stronger psychophysiological reactions as well as higher valence, arousal and craving ratings than cues without context, with physical and social smoking contexts combined putatively yielding the best results. Furthermore, we hypothesized that presentation of a fully intact cigarette would produce a stronger cue response on the physiological and perceptual level than presentation of a fully consumed cigarette.

## Methods

### Setting and participants

Twenty-two (22) individuals with TUD (twelve men, ten women, average age 36.37 (±4.02) years, range 18-51, with at least moderate TUD according to DSM-5[20] were evaluated. Test persons were recruited at the Central Institute for Mental Health in Mannheim as well as at the Universities of Mannheim, Karlsruhe, and Landau, and via newspaper advertisements. All participants had been smoking 12 cigarettes or more per day for at least one year. Severity of nicotine dependence was assessed using the short form Fagerström-Test [21, 22]. Test subjects were asked not to smoke in the hour preceding the experiment. As compensation for their time and effort participants were each reimbursed €20. Participants were excluded if they had any mental disorder other than TUD according to DSM-5 or any other SUD, except for tobacco smoking, screened with Structured Clinical Interview (SKID-I) for DSM-IV [23] due to the unavailability of a German SKID for DSM-5 at the time of examination or had a history of neurological problems. All participants had to be medication-free for at least 1 week prior to the assessment. Ethical approval was obtained from the local review board and written informed consent was obtained from all subjects prior to participation. Because the present study is a non-interventional study, it was not classified as a clinical trial and thus no preregistration in an ICMJE-approved registry took place. The present study was carried out in accordance with the Declaration of Helsinki.

### Assessments

#### Cue reactivity paradigm

##### Materials

We employed color photographs, in which either a group of smoking individuals, a cigarette on its own, or a hand holding a cigarette was depicted. Most pictures were original photographs, while some were downloaded from the internet and others taken from a study by Grüsser, Heinz and Flor [24] and a study by Pauli and Mucha [25]. The pictures were sorted into different context categories (ten pictures per category) depicting different aspects of a smoking situation, including physical contexts (cigarette in bar versus in a neutral environment; not showing people) and social contexts (people smoking in pub versus people smoking in neutral environment) as well as different time points within the smoking ritual (beginning of smoke consumption versus middle and terminal stages of smoke consumption). As control conditions, smoking cues without a context and, in addition, a black screen with a white fixation cross was used (see Figure 1).

**Figure 1:**
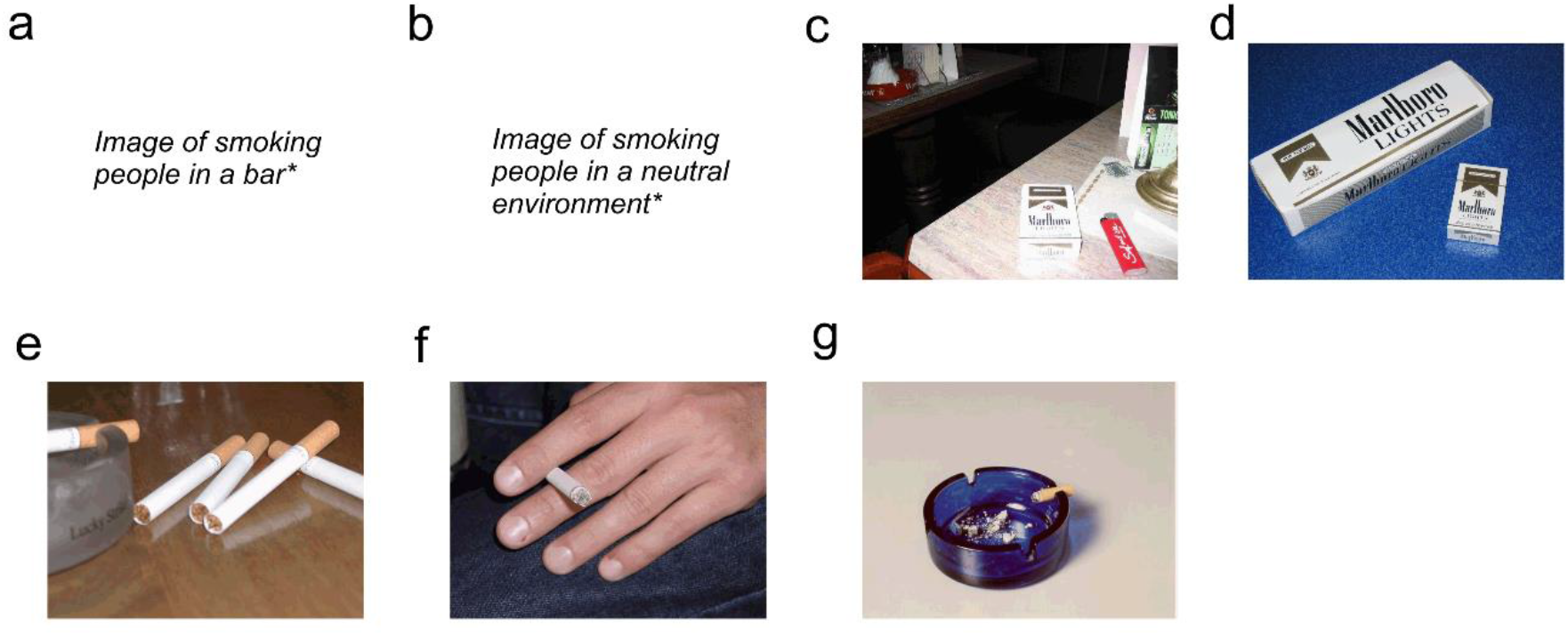
Visual stimulus material of physical (bar versus neutral environment) and social (people in bar versus people in neutral context) smoking contexts; different phases of the smoking-ritual (beginning, middle and terminal phase). One example for each category is given: (a) people smoking in bar (social, physical); (b) people smoking in neutral environment (social); (c) bar, no people (physical); (d) no context; (e) beginning phase; (f) middle phase; (g) terminal phase. *Due to publication guidelines the original pictures have been removed here.

Five pictures (one of each category including smoking cues) were presented during a short habituation phase. Each of the 70 pictures was presented once for 6 seconds on a high-resolution 21″ monitor (Iiyama Vision Master PRO 500, Iiyama International b.v., BC Oude Meer, the Netherlands) placed 1 m in front of the subject with an interstimulus interval of 7–12 seconds and in random order, different for each subject and with not more than two stimuli of the same category in series.

##### Ratings

After the assessment of cue reactivity, participants rated the emotional valence and arousal of each picture using the Self-Assessment-Manikin[26]. Subsequently, a short questionnaire of “like - dislike” questions with possible yes / no responses related to craving, typical situations and disliking was administered to assess participants’ perceived cigarette craving in response to the presentation of the single pictures. All pictures were presented in random order, different for each subject and different from the order during the psychophysiological assessment. The complexity of all pictures used was rated and counterbalanced. To check for validity, pictures were rated in a pre-study sample of 40 students.

##### Physiological recordings

The physiological data (EMG, SCR and ECG) were acquired with a Nihon-Kohden-Polygraph (Nihon Kohden Europe, GmbH, Rosbach, Germany).

##### Startle response modulation

To evoke the startle reflex, we used a white noise stimulus (95 dB/A, 50 ms) presented via headphones 1.5–3.5 seconds after picture onset. The first five startle probes - delivered after five test pictures displayed for 3 seconds - served as habituation trials and were not included in further analysis. The following startle probes were delivered during half of the 70 pictures of every category and five times during the fixation cross to minimize predictability and to be able to analyze skin conductance responses (SCRs) without interference of the startle response. The startle response was assessed as peak activity of the left musculus orbicularis oculi. The EMG signal was registered at a sampling rate of 1000 Hz with a low-pass filter of 500 Hz and a high-pass filter of 28 Hz. The raw EMG signals were rectified and integrated with a time constant of 20 ms. The startle response represents the difference between a stable baseline (using the 50 ms before picture onset) and the maximum amplitude of the startle reflex 50–150 ms after presentation of the startle probe. The startle response was standardized (T scores) using means and SDs across all stimulus categories and across all subjects to minimize between-participant variability in the absolute size of startle responses [50 + 10(x-mean)/SD]. In a visual analysis, all trials were inspected with respect to non-response (no visible startle response: < 5 mV) and artefacts (i.e. multiple peaks, voluntary or spontaneous eyeblinks at or near the startle stimulus onset)[27]. For data analysis, we used only data of participants who responded to the white noise stimulus in at least 75% of the trials without artefacts and had at least four valid trials per stimulus category. Data analysis was carried out using a custom-made, self-written program running on Matlab software (The Math Works Inc., Natick, MA, USA).

##### SCR

For the assessment of SCRs, electrodes were placed on the thenar and hypothenar eminence of the non-dominant hand and supplied with a constant voltage of 0.5 V. Skin conductance was registered at a sampling rate of 20 Hz, with a low-pass filter of 15 Hz and a time constant of 10 s. SCR parameters were derived using the software EDR_PAR (written by Dr. Florian Schäfer, Department of Physiological Psychology, University of Wuppertal, Germany). The SCR was assessed as the maximum amplitude 1–4 seconds after picture onset. SCR data below 0.01mS were classified as zero responses. Raw signals were log (1 + SCR) transformed to ensure a normal distribution of the data. SCR was only analyzed for pictures without startle probe presentation.

##### Heart rate

ECG was registered at a sampling rate of 1000 Hz, with a low-pass filter of 70 Hz and a time constant of 0.1 s. Data analysis was conducted using the software IBI_PARA (written by Dr. Florian Schäfer, Department of Physiological Psychology, University of Wuppertal, Germany). After R-wave detection, the deviation between mean heart rate during six-second picture presentation and three-second baseline was calculated.

### Data analysis

Ratings of emotional valence, arousal, craving as well as startle reflex magnitude, skin conductance and heart rate were analyzed with a repeated measures analysis of variance using a 2 (physical/no context) x 2 (social/ no context) x 3 (smoking-ritual: beginning versus middle versus terminal phase) factorial design. For the craving data (like-dislike) sumscores of the ‘yes’ answers were calculated. Furthermore, we tested for normal distribution of the data.

For all analyses, α was set to 0.05. In the case of violation of the assumption of homogeneity of variance, we applied Greenhouse–Geisser adjustments and report adjusted degrees of freedom. Planned paired t-tests with Bonferroni corrections were used to decompose significant main effects or interactions. For all statistical analyses, we used the Statistical Package of the Social Sciences (SPSS), Version 25.0.1 for Windows (SPSS Inc., Chicago, IL, USA).

## Results

### Psychophysiological assessments

#### Startle response modulation

A trend towards significance of the repeated measurement factor cue category (F(7, 147)=2.03, p=0.06) for the maximal amplitude of the blink reflex was observed (see Figure 2a). A comparison between the cues in social contexts and cues without contexts were not significant (F(2, 42)=0.97; p=0.39. However, a significant difference between spatial context cue categories and the comparison cue category ‘black screen’ was indeed noted (F(2, 42)=4.02; p=0.023). Blink reflex reactions during ‘black screen’ cue presentations differed significantly from those observed during ‘spatial context: bar’ image presentations (p=0.03), but not from those recorded during ‘spatial context: neutral’ image presentations (p=0.71). With regards to cigarette consumption status, no significant difference in the startle reflex could be determined across categories (F(3, 63)=1.00, p=0.40). Startle response latency was not significantly modulated by the categories (F(7, 147)=1.65, p=0.13) (see Figure 2b).

**Figure 2:**
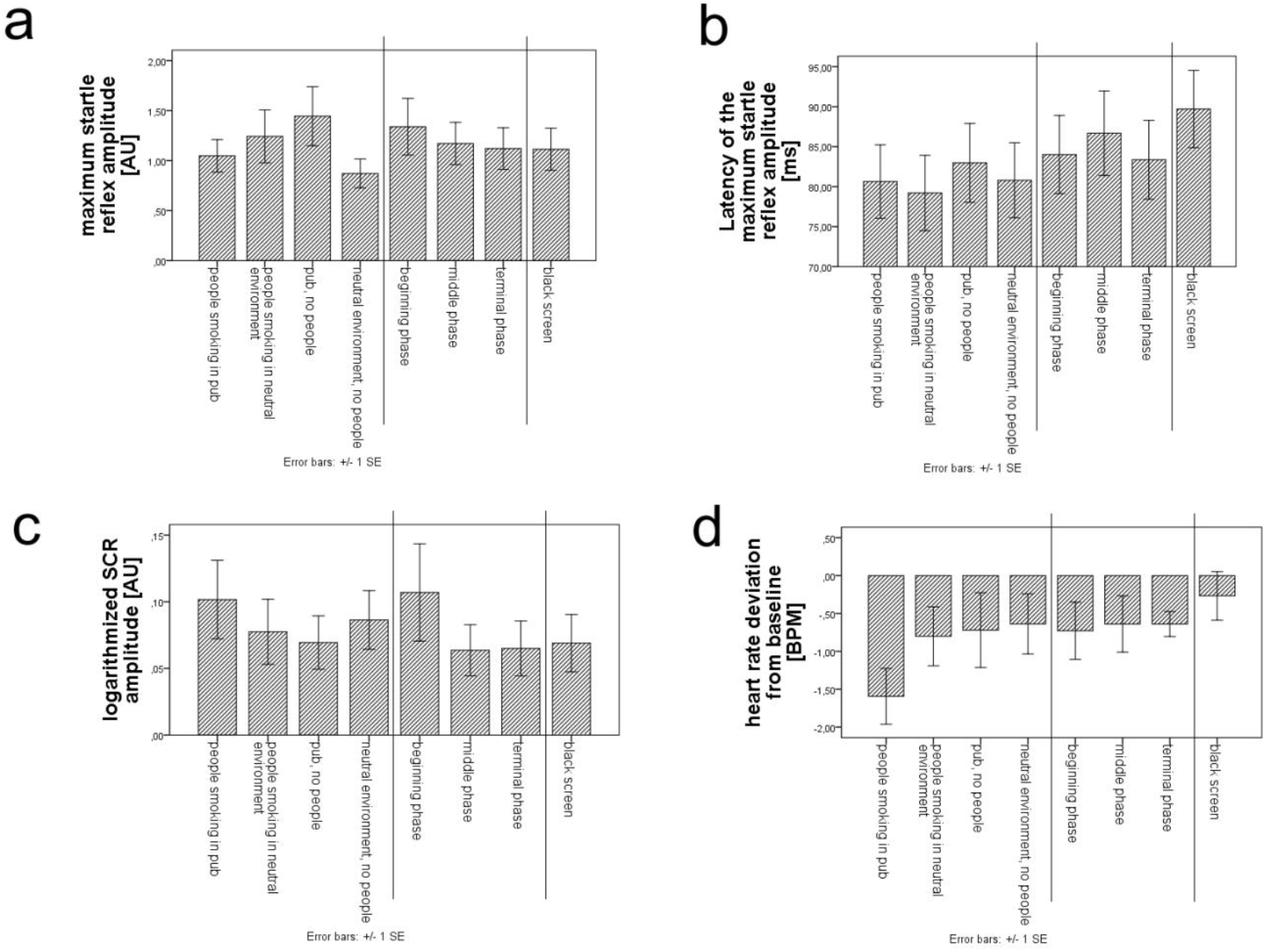
Mean (± SE) of the physiological responses evoked by different contexts; a) maximum startle reflex amplitude [arbitrary units] b) latency of the maximum startle reflex amplitude [milliseconds after stimulus onset] c) logarithmic SCR amplitude [arbitrary units]; d) average heart rate deviation from baseline [beats per minute, BPM]

#### SCR

Cue/context category was not significant for SCR (F(7,147)=0.83; p=0.57) (see Figure 2c). Comparisons within single cue categories did not reveal significant differences in phasic skin conductance responses for social (F(2, 42)=1.23; p=0.30) or cigarette consumption status (F(3, 63)=90; p=0.45).

#### Heart rate

Due to a technical artefact, only 14 of 22 total data sets could be included in the final analysis. The analysis of variance did not show any significant differences in heart rate change between cue/context categories (F(7,91)=1.23; p=0.28) (see Figure 2d). An analysis of variance for social context did, however, generate significant results (F(2,26)=3.64; p=0.04). Heart rate changes to the cue/context categories ‘social’ and ‘neutral’ differed significantly (p=0.037). The mean heart rate deceleration during presentation of ‘neutral smoking context’ image cues/contexts (M= −0.80) was smaller than the mean heart rate deceleration during ‘social smoking context’ image cue/context presentation (M=-1.60). Responses to the category ‘social smoking context’ also differed significantly. However, no significant difference was observed between responses to the comparison category and cue/context category ‘neutral smoking context’ (p=0.85). With regards to the two spatial context cue categories, no significant differences were determined (F(2, 26)=0.45; p=0.64). Finally, comparisons with all three cue categories of cigarette consumption status likewise produced no significant differences in mean change in heart rate (F(3, 39)=0.47; p=0.70).

The cue/context category ‘neutral smoking context’ resulted in a deceleration of heart rate up until one second post-image presentation. Thereafter, a steady acceleration in heart rate was recorded. Temporal analysis of heart rate in response to ‘social smoking context’ cue images revealed a pattern in which heart rate decelerated up until 3 seconds from the time of image presentation, followed by a subsequent acceleration. The comparison category ‘black screen’ initially resulted in heart rate acceleration. Heart rate responses to images in the cue categories ‘fully smoked’, ‘unsmoked’, and ‘black screen’ were characterized by a brief initial acceleration, followed by deceleration up until the third second, and a final switch back into an accelerating mode for the rest of the observation period (see supplementary Figure 1b). The heart rate time course for ‘half-smoked’ cue images is best described as initial deceleration followed by acceleration. Lastly, all spatial context cue categories followed a tripartite pattern of acceleration – deceleration – acceleration with regards to heart rate response (see supplementary Figure 1c).

### Craving questionnaires

#### Short questionnaire „like-dislike”

Participants perceived pictures belonging to the cue/context categories ‚social context‘, physical context’, and ‘consumption status: unsmoked cigarette’ as most salient for triggering craving (see supplementary Figure 2a). Pictures containing a social context elicited a greater degree of craving than pictures containing a neutral context. The ability of nicotine images to elicit craving also depended greatly on the spatial context within which the cigarette was presented. The context ‘Bar’ was also rated as significantly more craving-inducing than a neutral spatial context.

Images depicting social contexts (M = 6.36, SD 3.61; t = 5.477, p < .001, n = 22) were significantly more effective in craving-inducing than mages containing neutral contexts (M = 3.68, SD = 3.00). The neutral physical context (M = 3.68, SD = 3.00) reduced cigarette craving more than the physical (bar) context (M = 5.77, SD = 3.02; t = 3.828, p < .001, n = 22). Regarding the consumption status of a cigarette, images depicting unsmoked cigarettes (M = 5.50, SD = 2.76), when compared to images of fully smoked (M = 1.41, SD = 2.30) cigarettes, were significantly more craving-inducing, t = 6.042, p < .001, n = 22. However, the difference in feelings of craving between an unsmoked cigarette (M = 5.50, SD = 2.76) and a half-smoked cigarette (M = 5.18, SD = 3.33), were not nearly as pronounced, t = 0.405, p < .689, n = 22.

Images using a ‘social smoking context’, ‘cigarette consumption status: half-smoked’ and ‘spatial context: bar’ were judged as very typical situations (see supplementary Figure 2b). The categories ‘neutral context’ and ‘cigarette consumption status: fully smoked’, ‘cigarette consumption status: unsmoked’ and ‘spatial context: neutral’ were judged to be significantly less typical.

#### Valence and Arousal Assessments

Valence ratings were significantly different between the different categories of images (cue/context category: F(6, 126)=19.29; p<0.001) (see supplementary Figure 3a). Pairwise single comparisons revealed that the ‘social smoking context’ was perceived as marginally significantly more pleasant (M=5.98) than the ‘neutral smoking context’ (M=5.19, p=0.08). Here, images belonging to the category ‘social smoking context’ were rated as more pleasant. The two spatial cue/context categories (‘bar context’ and ‘neutral context’) were not rated as significantly different in their valence by study participants. The cue/context category ‘cigarette consumption status: fully smoked’ showed a significant difference with all of the other cue categories evaluated (p=0.001). As a result, this cue category was judged to be least valent.

For arousal ratings, we also found a significant difference in the average arousal scores between the individual cue/context categories (see supplementary Figure 3b, cue category; (F(6, 126)=3.22; p<0.05). Pairwise single comparisons between the cue categories ‘social smoking context’ and ‘neutral smoking context’ (p=0.103), between ‘social smoking context’ and ‘cigarette consumption status: half-smoked’ (p=0.164) as well as between ‘social smoking context’ and ‘cigarette consumption status: fully smoked’ (p=0.309) revealed no significant differences. Images of the category ‘social smoking context’ (M=4.54) were rated as more arousing than images in the ‘neutral smoking context’ (M=3.86) cue category. In evaluating the effect of spatial context (‘bar context’ and ‘neutral context’) and cigarette consumption status (‘unsmoked’, half-smoked’ and ‘fully smoked’), no significant differences in arousal between groups were found.

## Discussion

In the present study, valence arousal and craving ratings were assessed during different context categories in an cue reactivity paradigm and SCR amplitude, mean heart rate deviation from baseline, and maximal blink reflex amplitude were used as physiological correlates of cue-induced nicotine craving. Our main findings are a significantly stronger heart rate deceleration in the social versus neutral smoking context and an increased startle response in the physical context (bar) versus control (fixation cross) context. Contrary to our expectations, no significant differences in SCR were observed between the individual cue categories. Nicotine cues that were presented within a social context elicited a greater decrease in heart rate than those presented within a neutral context. A decrease in heart rate can be interpreted as the inability to reorient attention away from a preoccupying stimulus. Thus, the social smoking context appears to have been particularly attractive and motivating for study participants.

The ‘neutral smoking context’ and ‘social smoking context’ both caused a deceleration in heart rate, while the display of the black screen produced an acceleration. The deceleration represents an orienting response to the new stimulus, which generally occurs at the beginning of cue image presentation. The heart rate time course in response to ‘social smoking context’ cues provoked not only this standard initial deceleration, but a prolonged deceleration up until the third second of cue presentation. Following the third second, an acceleration was recorded. In contrast, the ‘neutral smoking context’ merely produced a deceleration lasting up to the first second, beyond which an acceleration was observed. This rapid switch to acceleration could be interpreted as the result of shifting attention away from the nicotine cues presented in the neutral environment. Thus, these cues may have been experienced as less attractive and stimulating by the participants. Interestingly, cues in the categories ‘spatial smoking context’ and ‘consumption status’ did not produce the orienting response. The spatial contexts (‘smoking in bar’ and ‘smoking in neutral environment’) instead demonstrated a triphasic heart rate time course, in which modulations followed a pattern of acceleration – deceleration – acceleration.

The analysis of startle reflex responses showed a trend towards significance with respect to the repeated measurement factor cue/context category. It was demonstrated that blink reflex reactions to the comparison category ‘black screen’ differed significantly when compared to reactions to nicotine cues presented within a bar context. Furthermore, maximal blinking reflex amplitudes were shown to have statistically occurred at the same point in time (average timing: 83 ms).

Nicotine cues presented in a social context produced a slowing of heart rate and presumably an increased craving to smoke when compared to nicotine cues presented in a neutral context. However, this effect was not observed using the SCR and EMG modalities to examine cue category differences. Moreover, the assumption that nicotine cues belonging to the category ‘spatial context: bar’ would elicit a greater psychophysiological response than those belonging to the category ‘spatial context: neutral’ was also not borne out. Although a comparison of the median of each of these cue categories indicated that smokers reacted more strongly to cues presented in the bar context, statistical significance was not achieved. Multiple explanations could account for this observation.

As previously reported by Avants, Margolin, Kosten, and Cooney [28], many test subjects do not react to these types of visual cues. In past studies, it was observed that roughly one third of all study participants did not show any psychophysiological responses to drug-relevant cues [6, 7]. Different factors that modulate reactivity to drug cues could be responsible for this finding. In particular, personality-dependent and SUD-related aspects, such as degree of the severity of the SUD or environmental stimuli/noise, which could distract attentional focus, could potentially account for non-response [9, 29]. This phenomenon, in which different individuals react more or less strongly to drug cues, could perhaps even partially explain the variance in individual susceptibility to TUD. Other factors contributing to individual variability in drug cue reactivity might include duration of smoking cessation and class of smoker type. Furthermore, it is possible that not all nicotine cues used in the present study acted successfully as conditioned cues and consequently did not generate any cigarette craving, as a result. From a methodological perspective, it is useful to use standardized visual nicotine cues across all subjects, although this naturally introduces the limitation that conditioning processes unique to each individual smoker are not taken into consideration. It is therefore possible that the present sample was too small to prevent systematic effects on the data resulting from presented cues not constituting relevant and salient nicotine cues for some participants.

We also assumed that a fully unconsumed cigarette would generate a more pronounced psychophysiological response (high SCR amplitude, significant deviation in average heart rate, high EMG amplitude) than presentation of a fully consumed cigarette, with half-smoked cigarettes in between. However, we found no significant differences in phasic skin conductance response, mean deviation in heart rate, and startle EMG amplitude between these categories. However, both fully smoked and unsmoked cigarette cues produced an initial acceleration in heart rate, while half-smoked cigarette cues did not. These nicotine cues appear to have been perceived as attractive by participants. It is possible that the artificial laboratory environment prevented participants from experiencing true feelings of craving in response to otherwise stimulating cues and therefore they showed no measurable psychophysiological responses. Perhaps a different result would have been achieved in more familiar smoking environments. Smokers experience cravings when their blood nicotine level sinks below a certain threshold, which varies between individuals. It is therefore also possible that the hour-long period of smoking abstinence enforced before the beginning of the study and the associated drop in blood nicotine level was not sufficient to elicit physiological changes not only in response to ‘social context’ nicotine cues, but also to nicotine cues consisting solely of the cigarette itself. Smokers for whom social context plays a role in driving craving are presumably not as severely nicotine dependent.

We also assumed that craving to smoke is most effectively triggered by those visual cues that combine a social smoking context with bar setting. Nicotine images depicting neutral situations or fully consumed cigarettes should not trigger any cravings, or even reduce cravings, acting as aversive stimuli. Regarding cue image reactivity, it would be expected that ratings of the presented images would differ from the results obtained via physiological measurements. Evaluating this assumption using ‘like-dislike’ questionnaires, it was observed that nicotine cues presented within a social smoking context were indeed rated as more craving-inducing than cues in a neutral context. Furthermore, images depicting fully unconsumed cigarettes were judged as being more enticing than images depicting fully smoked cigarettes, though only marginally more craving inducing than images of half-smoked cigarettes. The physical context ‘bar’ was rated as generating more desire to smoke than neutral physical contexts. Images containing fully smoked cigarettes were graded as most undesirable, reducing craving to smoke to the greatest extent. Taken together, these results support the high significance of context for craving.

For the emotional processing of nicotine cues, significant differences could be observed in the valence scores assigned to images of differing cue/context categories. Valence scores did not differ significantly between images containing nicotine cues consisting of half-consumed cigarettes vs. those consisting of unconsumed cigarettes. However, both of these cue categories did differ significantly from valence scores assigned to nicotine cues depicting fully consumed cigarettes. Images depicting half consumed cigarettes or unconsumed cigarettes were rated as significantly more pleasant than images showing fully consumed cigarettes. Images representing a social smoking context were not rated as more pleasant than those representing a neutral context. Furthermore, no significant difference in valence scores between the physical smoking contexts ‘bar’ and ‘neutral’ were found. Other significant differences in assigned valence scores could be observed between the unpaired cue categories ‘social smoking context’ and ‘consumption status: fully smoked’, and between ‘social smoking context’ and ‘neutral physical context’. Images involving social contexts were judged as more valent by participants. To sum up, images associated with the cue category ‘social smoking context’ were rated as most pleasant, while cues associated with the category ‘consumption status: unconsumed’ were rated as least pleasant.

In addition, significant differences in arousal scores became evident between differing cue categories. Images belonging to the cue category ‘social smoking context’ were perceived as more arousing than images belonging to the category ‘neutral smoking context’. For physical contexts, (‘drug in a bar context’ and ‘drug in a neutral context’) and consumption status (‘unconsumed’, ‘half-consumed’, ‘consumed’), no significant differences were observed within each category. The cue category ‘social smoking context’ also significantly differed from arousal ratings assigned to the categories ‘consumption status: half-consumed’ and ‘consumption status: consumed’. With respect to the reactivity of individual image cues, the hypothesis is supported in so far as different cues barely elicited a physiological response and, as a result, no major variation between cue categories could be observed, whereas the ratings of arousal and valence did significantly differ between the individual cue/context categories, as did self-reports of craving.

One possible explanation for the general lack of effect of the contexts on SCR might be the high variability of the data. Presumably, large interindividual differences heterogenize the data to an unmanageable degree. In other words, the large variance results in a mean without much explanatory power, resulting in the absence of any detectable statistical significance. Whether the variance is obscuring a real effect presently remains unclear.

### Clinical implications

Our findings indicate that social versus neutral smoking contexts and confrontation with smoking cues in bars are associated with enhanced cue-reactivity. These results have direct implications for treatment approaches aiming to reduce cue-reactivity like cue-exposure-based extinction treatment (CET). CET could be more efficacious if context is included, e.g. when the CET takes place in real bars or in laboratory settings with a bar environment (“BarLab”) and also in group therapy settings. Also cognitive behavioral therapy (CBT) including stimulus control strategies could include contexts, for example, patients should assess individual situations, thoughts and other contexts, which trigger craving with the aim to avoid and control them.

Further, implications can also be drawn for prevention. Tobacco advertisement including social contexts and bar environments might be more appealing than neutral contexts. This interpretation fits our findings that tobacco advertisement elicitsneural cue-reactivity even in non-smokers[29]. Prohibition of such advertisements might be an efficacious prevention strategy.

### Limitations

A potential limitation of our study is the fairly small sample size. Therefore, replication of our results in a larger sample is needed. This would, as a natural consequence, also increase statistical power.

## Data Availability

All data produced in the present study are available upon reasonable request to the authors

## Acknowledgments

The present study was supported by a grant from the Deutsche Forschungsgemeinschaft (TRR 265 Project ID-402170461, C01 SVK/HF). We would like to thank Tina Stonner for her contribution to data acquisition and data analysis. Furthermore, we would also like to thank Alycia Lee, B. Sc., for her contribution in translation and language-editing.

