## Supplementary figures and images for "Contexts enhance ratings of craving and psychophysiological responses of cue-reactivity in tobacco use disorder"

### Supplementary Figure 1

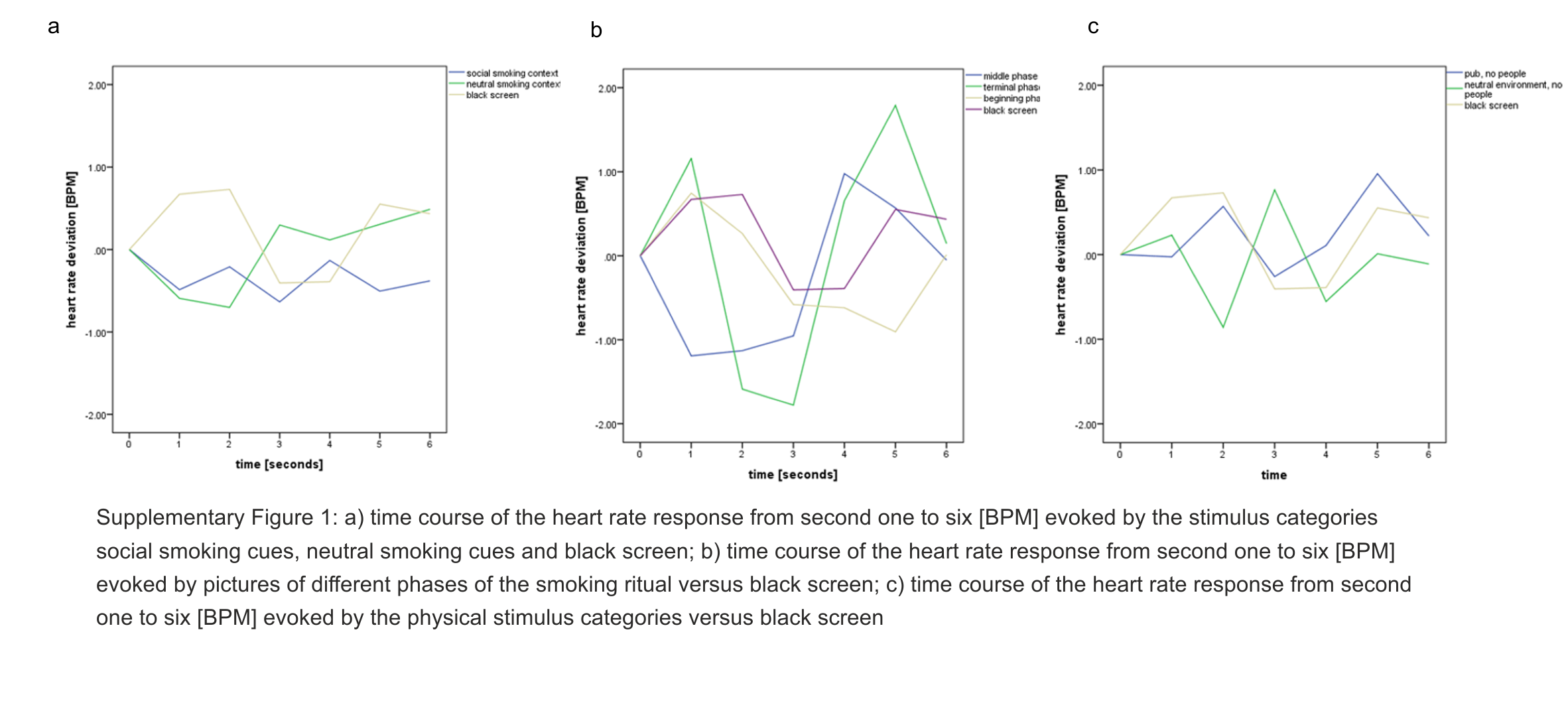

### Supplementary Figure 2

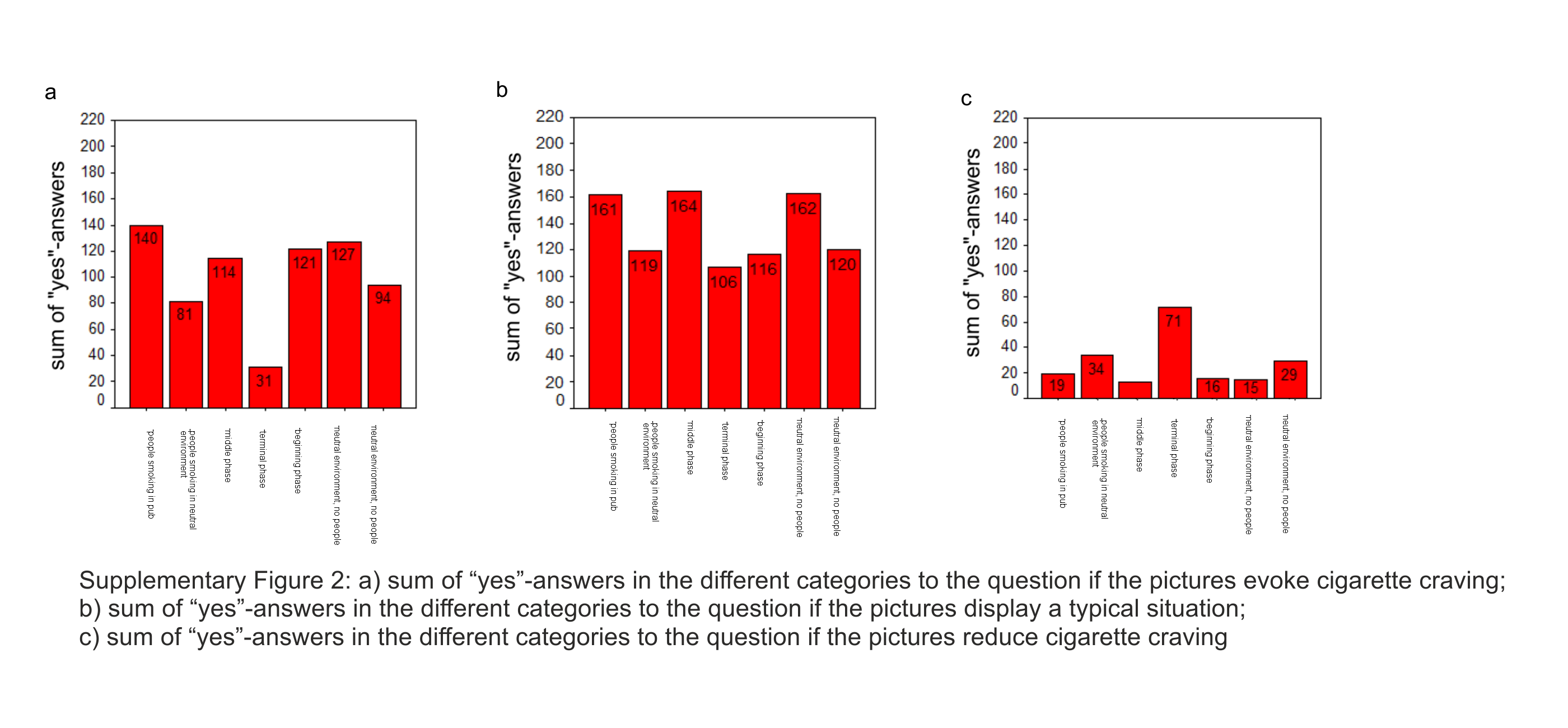

### Supplementary Figure 3

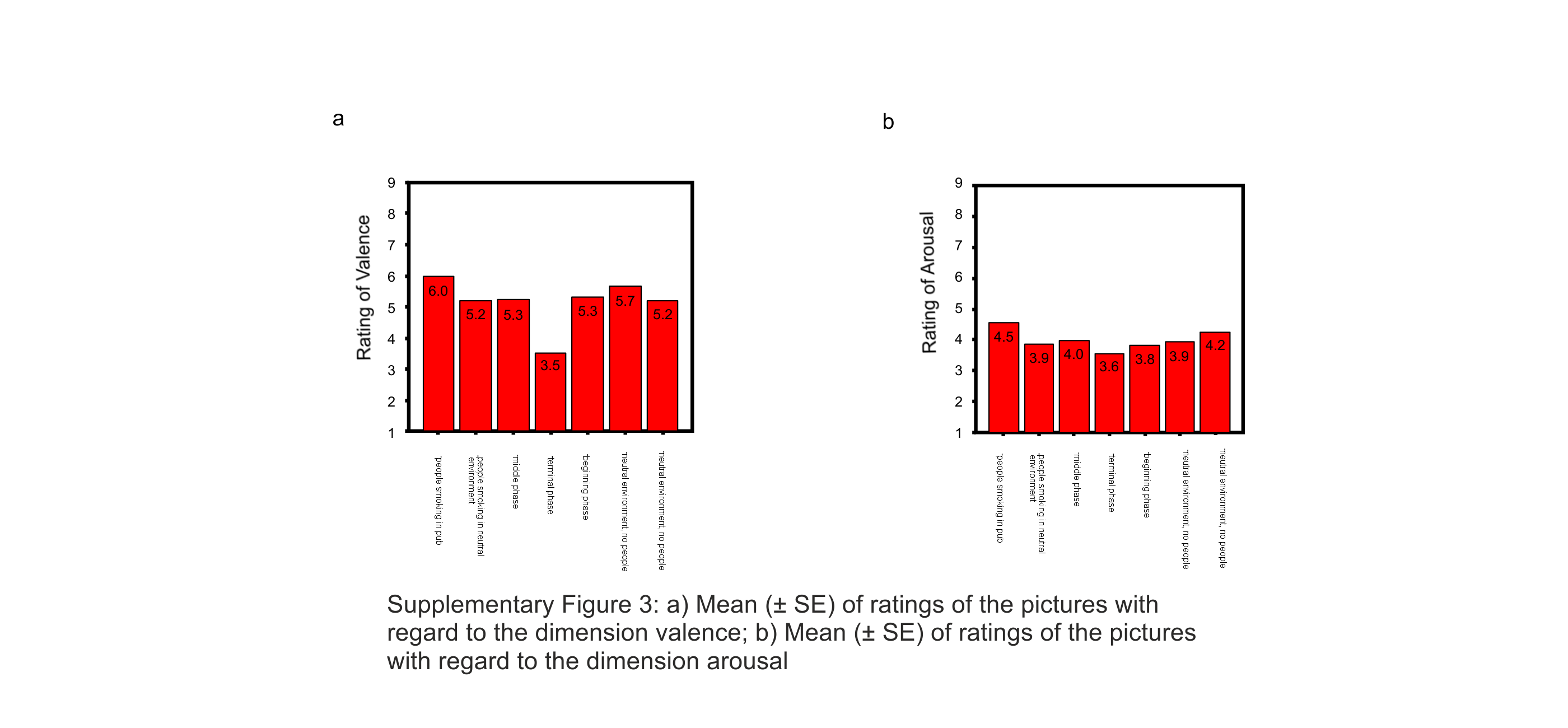
